# Seasonal purchase of antihistamines and ovarian cancer risk in the Cancer Loyalty Card Study (CLOCS): results from an observational case-control study

**DOI:** 10.1101/2023.05.30.23290729

**Authors:** Hannah R. Brewer, Qianhui Jiang, Sudha Sundar, Yasemin Hirst, James M. Flanagan

**Author notes:** **Address for correspondence:** Dr. James M. Flanagan, Division of Cancer, Department of Surgery and Cancer, Faculty of Medicine, Imperial College London, 4th Floor IRDB, Hammersmith Campus, Du Cane Road, London W12 0NN.

## Abstract

**Objective:** Antihistamine use has previously been associated with a reduction in incidence of ovarian cancer, particularly in pre-menopausal women. Herein, we investigate antihistamine exposure in relation to ovarian cancer risk using a novel data resource by examining purchase histories from retailer loyalty card data.

**Study Design:** A subset of participants from the Cancer Loyalty Card Study (CLOCS) for which purchase histories were available were analysed in this study. Cases (n=153) were women in the UK with a first diagnosis of ovarian cancer between Jan 2018 – Jan 2022. Controls (n=120) were women in the UK without a diagnosis of ovarian cancer. Up to 6 years of purchase history was retrieved from two participating high street retailers from 2014-2022.

**Main outcome measures:** Logistic regression was used to estimate the odds ratio (OR) and 95% confidence intervals (CIs) for ovarian cancer associated with antihistamine purchases, ever versus never, adjusting for age and oral contraceptive use. The association was stratified by season of purchase, age over and under 50 years, ovarian cancer histology, and family history.

**Results:** Ever purchasing antihistamines was not significantly associated with ovarian cancer overall in this small study (OR:0.68, 95% CI: 0.39,1.19). However, antihistamine purchases were significantly associated with reduced ovarian cancer risk when purchased only in spring and/or summer (OR: 0.37, 95% CI: 0.17,0.82) compared with purchasing all year (OR: 0.99, 95% CI: 0.51,1.92). In the stratified analysis, the association was strongest in non-serous ovarian cancer (OR: 0.41, 95% CI:0.18,0.93).

**Conclusions:** Antihistamine purchase is associated with reduced ovarian cancer risk when purchased seasonally in spring and summer. However, larger studies and more research is required to understand the mechanisms of reduced ovarian cancer risk related to seasonal purchases of antihistamines and allergies.

## Introduction

Ovarian cancer is the most lethal gynaecological disease around the world [1]. Due to its nonspecific symptoms and lack of screening tools, more than 70% of women are diagnosed when ovarian cancer has already progressed to the late stage, making it difficult to give patients a prompt and effective treatment [1, 2]. The five-year survival rate of women with advanced ovarian cancer is less than 20%, while women diagnosed with stage I have more than 90% chance of five-year survival [3]. Currently in the UK, population-based screening for ovarian cancer is not recommended [4]. However, risk stratified screening or symptomatic screening remain viable options [5, 6].

A recent report suggested that antihistamine use may be associated with reduced ovarian cancer risk [7]. In a population-based case-control study investigating prescribed antihistamines in ovarian cancer patients (n=5,556) and controls (n=83,340), there was no overall association between ever use of antihistamines and ovarian cancer risk (OR:⍰0.97,95% CI: 0.90,1.05). However, in a subgroup analysis it appeared to be protective in women younger than 50 years (OR:⍰0.72, 95% CI:⍰0.57,0.90) and for mucinous ovarian cancer, specifically (OR:⍰0.74, 95% CI:⍰0.57,0.96).

There is a potential mechanistic basis for considering antihistamines as cancer preventive agents. Histamine is a biogenic amine synthesized from histidine, an essential amino acid, and catalysed by the L-histidine decarboxylase enzyme. After it is synthesized, histamine is stored in various cells, mainly in mast cells [8]. After being released, histamine can participate in cell proliferation and differentiation, inflammation, vasodilation, and neurotransmission [9, 10]. Notably, histamine also attenuates the immune response during immunotherapy, and antihistamines can reverse this effect [11]. This implies that histamine may play an essential role in tumorigenesis and progression. Hence, antihistamines that block the activity of histamine may have the opposite effect by blocking tumorigenesis. Alternatively, the reason many individuals use antihistamines is to treat allergies, which have also been suggested as a mechanism of cancer prevention [12, 13]. Therefore, it is important to further investigate the role of antihistamines and allergies in ovarian cancer prevention.

The Cancer Loyalty Card Study (CLOCS) is an observational case-control study aiming to investigate transactional data from retailer loyalty card programmes to help diagnose ovarian cancer earlier [14]. The main purpose of the CLOCS project was to examine the hypothesis that ovarian cancer patients purchase over-the-counter medications to manage certain symptoms in the 12 to 24 months before being diagnosed with ovarian cancer, i.e., there might be significant differences in purchase behaviour between ovarian cancer patients and individuals without ovarian cancer [14, 15]. In the present study, we hypothesised that ovarian cancer patients may be purchasing fewer antihistamines than control participants if it is a protective risk factor over the six years for which we have data available. Therefore, we aimed to investigate antihistamine purchases in relation to ovarian cancer risk.

## Methods

### Study Design, Setting, Participants

The CLOCS case-control study design and protocol have been described in detail previously [14, 15]. In brief, ovarian cancer patients diagnosed between 1^st^ January 2018 and 31^st^ January 2022 and recruited in 12 NHS clinic sites in England, Wales, and Scotland were considered cases. Any type of epithelial ovarian cancer was considered eligible (including high-grade serous, low-grade serous, endometrioid, clear cell, mucinous, borderline and other subtypes). Women without ovarian cancer were considered controls and were recruited online via the study website. Women living in the UK, 18 years or older, who owned at least one the participating retailers’ loyalty cards were eligible to take part. Participants with a previous cancer diagnosis and control participants who did not provide identity verification documents were excluded from analysis. After obtaining explicit consent, up to six years of loyalty card purchase history, ending at the date of recruitment, was requested from two high street retailers, referred to from here on as High Street Retailer 1 (HSR1) and High Street Retailer 2 (HSR2). One of the retailers offers a supply of health and beauty items, while the other stocks a wider range of health, beauty and grocery items.

### Variables

Participants completed a 24-Item Ovarian Cancer Risk questionnaire including key demographic characteristics (ethnicity, marital status, age [in years]) and other established risk factors including body mass index (BMI), age at menarche, menopausal status, age at menopause, parity, breastfeeding, hysterectomy, tubal ligation, cancer history, endometriosis, aspirin use, oral contraceptive (OC) use, hormone replacement therapy (HRT) use, family history of ovarian and breast cancers, vaping, and cigarette smoking. For cases, the date of diagnosis, histologic type, stage, grade, BRCA1 and BRCA2 status, and any surgical outcomes were completed by the clinical team. Pathology was confirmed by the recruiting clinical teams.

Antihistamine purchases were classified in the datasets from both retailers under the category of “Hayfever” including predominately cetirizine or loratadine tablets and other less common formulations. Purchases in cases were censored at the date of diagnosis.

### Statistical methods

A student’s t-test was used to test for a difference in age (continuous), and a chi-squared test was used to test for significant differences in ethnicity, loyalty card ownership, oral contraceptive use, and antihistamine purchases (categorical variables). Unconditional logistic regression was used to calculate risk of ovarian cancer among CLOCS participants unadjusted firstly, and then with adjustment for age and OC use (ever/never). Formal confounder testing was completed previously with all variables and found only age and OC use as significant confounders of case-control status in this dataset [15]. An additional sensitivity analysis was conducted with and without adjusting for store card ownership (HSR1, HSR2, or both HSR1 and HSR2) and in individual years prior to diagnosis, however, the results remained unchanged (data not shown). All statistical analyses were conducted in the Secure Enclave environment at Imperial College London using R (version 4.1.2).

## Results

The characteristics of all CLOCS participants have been described in detail elsewhere [15]. The characteristics for 153 participants with ovarian cancer (cases) and 120 participants without ovarian cancer (controls), for whom purchase histories were available, are shown in Table 1. This study size was limited by the feasibility of recruiting participants to this study {Brewer et al BMJ Open in press}. The mean age of cases was higher at 64.5 years than controls at 52.8 years (Student’s t-test, p<0.001), and OC use was significantly lower in cases (75%) compared with controls (90%, p=0.004). In total, 106 participants had ever bought antihistamines, while 177 had never bought antihistamines, and there was a significant difference in antihistamine purchases between cases and controls (p=0.013) (Table 1). The dates of antihistamine purchases ranged from September 2014 to September 2021. However, most purchases in this study were between autumn 2016 and winter 2020 (Figure 1A). There were 356, 286, 148 and 134 purchases of antihistamines among all participants in spring, summer, autumn and winter, respectively (Figure 1B).

**Table 1:** Characteristics of CLOCS participants including age, ethnicity, loyalty card ownership, oral contraceptive pill use and antihistamine purchase history.

| Characteristic | Cases (n = 153) |  |  | Controls (n = 120) |  |  | P value<sup>2</sup> |
| --- | --- | --- | --- | --- | --- | --- | --- |
|  | Mean (SD<sup>1</sup>) | N | % | Mean (SD<sup>1</sup>) | N | % |  |
| **Age - years** | 64.5 (11.1) |  |  | 52.8 (12.1) |  |  | <0.001 |
| **Ethnicity** |  |  |  |  |  |  |  |
| White |  | 139 | 90.8 |  | 103 | 85.8 | 0.756 |
| Non-White |  | 13 | 8.5 |  | 12 | 10.0 |  |
| missing |  | 1 | 0.7 |  | 5 | 4.2 |  |
| **Loyalty card** |  |  |  |  |  |  |  |
| HSR1 card only |  | 40 | 26.1 |  | 49 | 40.8 | 0.032 |
| HSR2 card only |  | 37 | 24.2 |  | 26 | 21.7 |  |
| Both HSR1 and HSR2 |  | 76 | 49.7 |  | 45 | 37.5 |  |
| **Oral contraceptive pill use** |  |  |  |  |  |  |  |
| Never |  | 37 | 24.2 |  | 12 | 10.0 | 0.004 |
| Ever |  | 115 | 75.2 |  | 108 | 90.0 |  |
| **Antihistamine purchase** |  |  |  |  |  |  |  |
| Never |  | 104 | 68.0 |  | 63 | 52.5 | 0.013 |
| Ever |  | 49 | 32.0 |  | 57 | 47.5 |  |

<sup>1</sup>Standard deviation. <sup>2</sup>Statistical test was Student's t-Test for Age and Chi-squared test for categorical variables.

**Figure 1.**
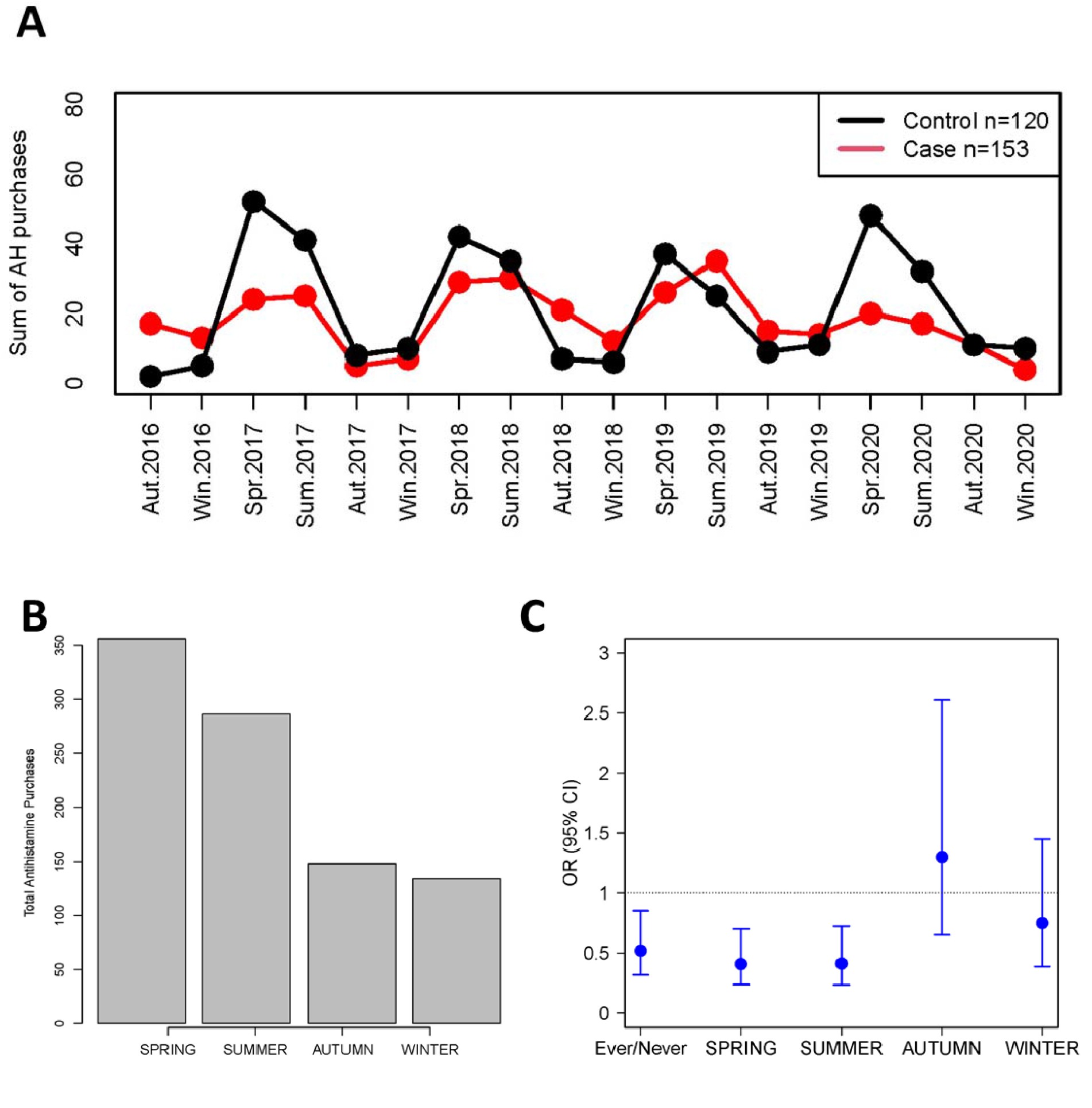
Antihistamine purchases in cases and controls. **A)** Purchases of antihistamines were summed across both retailers for each season from autumn 2016 to winter 2020 for cases (red, n=153) and controls (black, n=120). Seasons were defined as Spring (March, April, May), Summer (June, July, August), Autumn (September, October, November), and Winter (December, January, February). **B)** Total purchases for each season across all available data. **C)** Univariate logistic regression was performed for all purchases, and during each season as the predictor and case control status as the outcome. Odds ratios (Ors) are presented with error bars representing 95% confidence intervals (CIs).

In univariate analyses, there were significant associations with ovarian cancer risk for ever/never purchase of antihistamines, in spring and summer, but not in autumn or winter (Figure 1C, Table 2). After adjusting for age and OC use, ever purchase of antihistamines was not significantly associated with ovarian cancer with an adjusted OR of 0.68 (95% CI: 0.39,1.19) (Table 2). When stratified by season of purchase, there was a significant association in spring (OR: 0.52, 95% CI: 0.29,0.94) and summer (OR: 0.51, 95% CI: 0.28,0.95), but not in autumn or winter (Table 2). Stratifying the subjects into those that only bought antihistamines seasonally (in spring and/or summer) versus those who bought them all year (including winter and/or autumn) found that seasonal purchases were associated (OR: 0.37, 95% CI: 0.17,0.82) while purchasing all year was not (OR: 0.99, 95% CI: 0.51,1.92) compared with those who never bought antihistamines. In stratified analyses, there were null associations for participants over and under the age of 50 years, and family history of breast or ovarian cancer in a first-degree female relative. The association with ovarian cancer was stronger in non-serous ovarian cancer (OR: 0.41, 95% CI: 0.18,0.93) compared with serous ovarian cancer (OR: 0.90, 95% CI: 0.47,1.72).

**Table 2.**
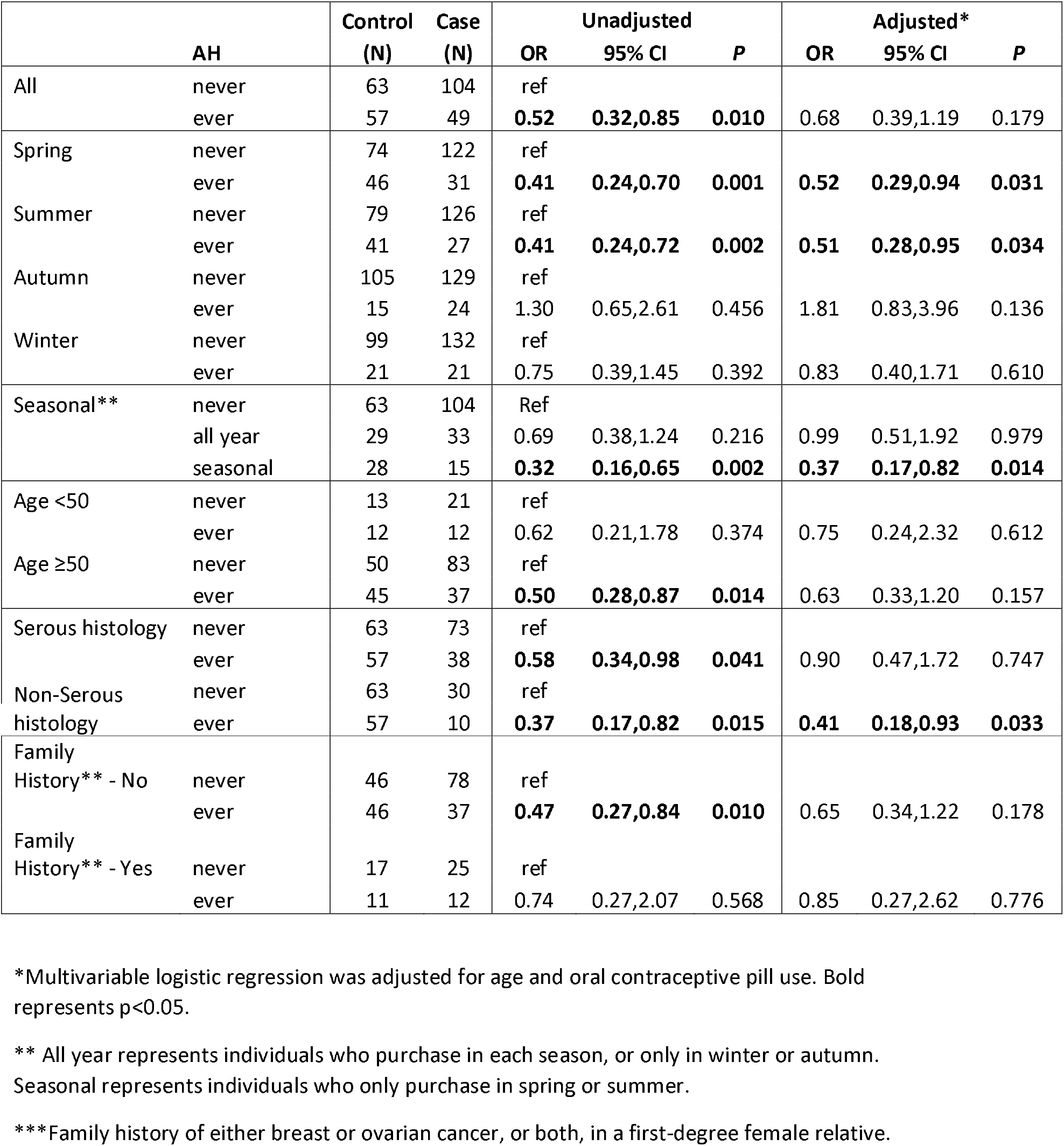
Purchase of Antihistamines (AH) and Ovarian Cancer Risk

|  | AH | Control<br>(N) | Case<br>(N) | Unadjusted |  |  | Adjusted* |  |  |
| --- | --- | --- | --- | --- | --- | --- | --- | --- | --- |
|  |  |  |  | OR | 95% CI | P | OR | 95% CI | P |
| All | never | 63 | 104 | ref |  |  |  |  |  |
|  | ever | 57 | 49 | **0.52** | **0.32,0.85** | **0.010** | 0.68 | 0.39,1.19 | 0.179 |
| Spring | never | 74 | 122 | ref |  |  |  |  |  |
|  | ever | 46 | 31 | **0.41** | **0.24,0.70** | **0.001** | **0.52** | **0.29,0.94** | **0.031** |
| Summer | never | 79 | 126 | ref |  |  |  |  |  |
|  | ever | 41 | 27 | **0.41** | **0.24,0.72** | **0.002** | **0.51** | **0.28,0.95** | **0.034** |
| Autumn | never | 105 | 129 | ref |  |  |  |  |  |
|  | ever | 15 | 24 | 1.30 | 0.65,2.61 | 0.456 | 1.81 | 0.83,3.96 | 0.136 |
| Winter | never | 99 | 132 | ref |  |  |  |  |  |
|  | ever | 21 | 21 | 0.75 | 0.39,1.45 | 0.392 | 0.83 | 0.40,1.71 | 0.610 |
| Seasonal** | never | 63 | 104 | Ref |  |  |  |  |  |
|  | all year | 29 | 33 | 0.69 | 0.38,1.24 | 0.216 | 0.99 | 0.51,1.92 | 0.979 |
|  | seasonal | 28 | 15 | **0.32** | **0.16,0.65** | **0.002** | **0.37** | **0.17,0.82** | **0.014** |
| Age <50 | never | 13 | 21 | ref |  |  |  |  |  |
|  | ever | 12 | 12 | 0.62 | 0.21,1.78 | 0.374 | 0.75 | 0.24,2.32 | 0.612 |
| Age ≥50 | never | 50 | 83 | ref |  |  |  |  |  |
|  | ever | 45 | 37 | **0.50** | **0.28,0.87** | **0.014** | 0.63 | 0.33,1.20 | 0.157 |
| Serous histology | never | 63 | 73 | ref |  |  |  |  |  |
|  | ever | 57 | 38 | **0.58** | **0.34,0.98** | **0.041** | 0.90 | 0.47,1.72 | 0.747 |
| Non-Serous histology | never | 63 | 30 | ref |  |  |  |  |  |
|  | ever | 57 | 10 | **0.37** | **0.17,0.82** | **0.015** | **0.41** | **0.18,0.93** | **0.033** |
| Family History** - No | never | 46 | 78 | ref |  |  |  |  |  |
|  | ever | 46 | 37 | **0.47** | **0.27,0.84** | **0.010** | 0.65 | 0.34,1.22 | 0.178 |
| Family History** - Yes | never | 17 | 25 | ref |  |  |  |  |  |
|  | ever | 11 | 12 | 0.74 | 0.27,2.07 | 0.568 | 0.85 | 0.27,2.62 | 0.776 |

\*Multivariable logistic regression was adjusted for age and oral contraceptive pill use. Bold represents $p < 0.05$ .
\*\* All year represents individuals who purchase in each season, or only in winter or autumn. Seasonal represents individuals who only purchase in spring or summer.
\*\*\*Family history of either breast or ovarian cancer, or both, in a first-degree female relative.

## Discussion

The Cancer Loyalty Card Study (CLOCS) is the first study to use transactional data from high street retailers to investigate cancer outcomes. While the main objective of CLOCS is to identify symptomatic purchases as an early diagnostic signal, we have focused in this study on cancer risk associated with specific purchase of antihistamines. A previous study reported an inverse association with prescribed antihistamine use and ovarian cancer in pre-menopausal women and specifically in the mucinous histological subtype [7]. Here, we have found an inverse relationship between antihistamine purchase and ovarian cancer risk when the purchases occur in spring and summer. In stratified analysis we found an association with non-serous ovarian cancer. Among CLOCS participants, mucinous ovarian cancer represented 12% of the non-serous cases, with endometrioid (29%) and clear cell (38%) making up the majority.

The main strength of this study is in the granular nature of the transactional data covering specific times, item descriptions, and doses of purchases that could allow in-depth analysis of specific products. With over 1.66 million transactions across 311 individuals over 7 years, there is a considerable depth to the available data. As a result, recall bias was avoided as a true picture of antihistamine purchases was obtained directly from retailers’ loyalty card programmes, which recorded in detail when antihistamines were purchased, all prior to the participants enrolling in CLOCS.

However, the main limitation of this study is that the purchase of antihistamines does not necessarily equate to the prolonged use of antihistamines. It is possible that people may be buying them for other reasons such as acute allergic reactions, not necessarily because they are experiencing seasonal allergy symptoms. Alternatively, they could be buying them for other household members or at other shops that were not included in this dataset. Other limitations include the small sample size and the age difference between the cases and controls, which was adjusted for in the multivariable analyses. There remains the possibility of additional confounding from unmeasured variables and other biases inherent in this study design. For example, the self-selection bias in the control recruitment may select for a more health-conscious population. There may be concern about the lag period between the purchases and onset of ovarian cancer (less than 6 years), however, it is possible that the pattern of purchases observed over those 6 years might accurately reflect the decades prior. Lastly, the COVID-19 pandemic occurred during recruitment which may have modified purchasing behaviour from March 2020 onwards. Fortunately, the data available in this study from 2014-2019 meant that we could investigate purchase patterns prior to the onset of the pandemic.

The previous study by Verdoodt and colleagues reported that women less than 50 years old showed the strongest protective effect with antihistamine use (OR = 0.72, 95% CI: 0.57,0.90) compared with women over 50 years (OR: 1.02, 95% CI:0.93, 1.11) [7]. In contrast, there was no significant difference between ages over and under 50 years among CLOCS participants, which may be explained by the smaller numbers of participants below the age of 50 years (n=25 cases vs n=33 controls). With small numbers we were also unable to stratify by individual histology subtype to explore the previously reported association with mucinous subtype.

One of the most surprising findings in this study was that seasonal purchases are associated with ovarian cancer risk. A trivial explanation for this finding may be that the higher numbers of purchases, increased during spring and summer, lead to stronger statistical power. However, it may also provide insight into the mechanism of the association with ovarian cancer risk. The predominant reason for purchasing antihistamines, during spring and summer, is allergies (e.g. seasonal allergies), which are suggested to prevent some cancers by promoting immune surveillance [12, 13]. This provides an example of “confounding by indication” meaning that the antihistamine purchase is a proxy for the real exposure of allergies. Some cancers in particular have a strong protective effect associated with allergies such as pancreatic cancer [16-18], lung [19], and head and neck cancers [20], but no effect on common cancers such as breast [19], prostate [19] and colon cancer [21]. The association reported for ovarian cancer with allergies was not significant (OR =0.75 (0.36-1.54)), with a small study size (n=38 cases), indicating large study sizes are required [13]. While the previous study [7] and ours did not have specific information about allergies from participants, collecting this information in future studies is warranted.

In summary, this study suggests that the use of antihistamines may have a protective effect against ovarian cancer, but whether this protective effect is robust needs to be investigated further. The mechanisms underlying how antihistamines reduce the risk of ovarian cancer also remains to be understood.

## Data Availability

Access to all Cancer Loyalty Card Study (CLOCS) purchase history data collected from the retailers is restricted due to the data sharing agreements in place and the sensitive nature of the data.

## Acknowledgments

The authors would like to acknowledge all the participants in CLOCS, the participating retailers, and the NHS sites that recruited ovarian cancer patients to this study. We would like to acknowledge Deborah Tanner and Fiona Murphy who have acted as ovarian cancer patient representatives since the inception of the CLOCS project. This study was funded by Cancer Research UK Early Detection and Diagnosis project grant (C38463/A26726) with additional support from the Cancer Research UK Imperial Centre, the National Institute for Health Research Imperial Biomedical Research Centre and the Ovarian Cancer Action Research Centre. The authors have no conflicts of interest to declare.

## Authors Contribution Statement

JMF, YH, HRB conceived the study and contributed to funding acquisition. HRB and JMF contributed to data curation. HRB, QJ and JMF conducted all formal statistical analyses. HRB, JMF and YH reviewed the results. JMF wrote the first draft of the manuscript. YH, HRB, QJ and SS commented and reviewed the final draft of the manuscript. All authors agree on the final version of the manuscript.

